# Biomedical Prevention without Social Protection: The ethics of providing PrEP to socially vulnerable adolescent girls and young women in rural Lesotho

**DOI:** 10.64898/2026.07.28.26358955

**Authors:** Andréa Williams, Tamaryn Crankshaw, Michael Strauss, Moruthoane Motlalentoa, Limakatso Mofilikoane, Mosa Mohasoa, Niklaus Labhardt

## Abstract

**Background:** Adolescence is a critical period for HIV prevention, yet many adolescent girls and young women (AGYW) experience this developmental transition within contexts of poverty, gender inequality, and limited social protection. As HIV pre-exposure prophylaxis (PrEP) expands across eastern and southern Africa, little attention has been paid to the ethical implications of providing PrEP within these social contexts.

**Methods:** We conducted a qualitative study in two rural districts of Lesotho using focus group discussions with AGYW aged 16–24 years and group interviews and individual interviews with village health workers and village chiefs. Data were analysed using reflexive thematic analysis.

**Results:** Three interconnected themes described how social vulnerability shaped young women’s sexual and reproductive health (SRH) and opportunities to benefit from PrEP. First, adolescence was characterised by limited SRH support, age-disparate and transactional relationships, and multiple forms of sexual violence occurring across homes, schools, hostels, and communities. Second, early and unintended pregnancy and child marriage accelerated transitions into adult roles and responsibilities before many young women were developmentally and socially prepared. Third, families, schools, communities, and existing protection systems frequently failed to provide adequate support, leaving young women to navigate violence, pregnancy, and early marriage with limited protection or recourse.

**Conclusions:** Providing PrEP to AGYW in contexts where social protections remain limited raises important ethical considerations. Expanding access to biomedical HIV prevention is necessary but insufficient. Ethical PrEP provision requires implementing social protection alongside biomedical prevention so that young women are supported not only to access PrEP, but also to meaningfully benefit from it.

*Highlights:*

- Adolescent girls experienced profound social vulnerability during critical developmental transitions.
- Early pregnancy and child marriage accelerated transitions into unsupported adult roles.
- Families and institutions frequently failed to provide adequate social protection.
- Ethical PrEP provision requires strengthening social protection alongside biomedical prevention.

## Introduction

HIV pre-exposure prophylaxis (PrEP) has transformed opportunities for HIV prevention among adolescent girls and young women (AGYW) by offering biological protection in contexts where unequal gendered relationships limit their ability to negotiate safer sex (Bekker et al., 2015; Delany-Moretlwe et al., 2022). PrEP has the potential to increase young women’s control over HIV prevention by reducing reliance on male partner cooperation for condom use (Dellar et al., 2015). While the use of PrEP is biologically protective, HIV vulnerabilities are oftentimes socially produced through poverty, unequal gender and age power dynamics, harmful social norms, and stigma, all of which constrain young women’s agency to protect themselves from HIV and engage with prevention services (Birdthistle et al., 2019; Pettifor et al., 2018; Stoebenau et al., 2016). Providing PrEP in these contexts raises important ethical questions about the responsibilities of the State to reduce HIV vulnerability and support young women’s engagement with HIV prevention.

Offering PrEP to AGYW is inherently ethically complex. During adolescence, young people are still developing autonomy and becoming more independent, yet they continue to rely on families, communities, and institutions for support and guidance as they make decisions about their SRH (Day et al., 2020; MacDonald et al., 2023; Patton et al., 2016; Sawyer et al., 2012). Yet in many high HIV burden settings, the social environments in which these developmental transitions occur are characterised by unequal gender norms, economic disadvantage, and limited SRH (SRH) education (Mukokoma et al., 2026; Stoebenau et al., 2016; Underwood et al., 2011). Ethical discussions surrounding PrEP provision for adolescents have largely focused on questions of distributive justic, equitable access, adolescents’ capacity to provide informed consent, confidentiality, and the long-term implications of PrEP use (Strode & Slack, 2015). This literature has mostly been developed in the context of people belonging to key populations living in high-income or well-resourced settings, with less attention to what ethical PrEP provision requires in contexts where vulnerability is socially produced and opportunities to exercise agency are structurally constrained. This shifts the ethical focus towards whether providing biomedical HIV prevention without broader social protections is ethically justifiable in contexts where AGYW experience intersecting vulnerabilities.

Lesotho provides an important setting in which to examine these questions. Despite substantial commitments to expanding HIV prevention, Lesotho has one of the highest HIV burdens globally, with AGYW disproportionately affected (*UNAIDS, 2025*). HIV prevalence among young women aged 20–24 years is four times higher than among young men of the same age (16.7% vs. 4.0%) (LePHIA, 2018). More than half of households live below the national poverty line, and the majority of the population resides in rural areas (LePHIA, 2018). Biomedical HIV prevention in Lesotho has expanded through community-based HIV services, differentiated models of care, and national PrEP guidelines, which allow adolescents aged 12 years and older to independently consent to HIV testing and HIV prevention services, including PrEP, without parental permission (Ministry of Health Lesotho, 2022). These policies create an enabling framework for adolescent PrEP provision.

However, supportive policies alone do not ensure that AGYW are able to meaningfully access, engage with, and benefit from biomedical HIV prevention. Rather, opportunities to engage with PrEP remain embedded within the social, economic, institutional, and geographic conditions of young women’s lives.

To better understand what ethical PrEP provision requires in this context, we conducted a qualitative study among AGYW aged 16–24 years living in rural Lesotho. Through focus group discussions with AGYW and VHWs, and in-depth interviews with village chiefs, we explored young women’s experiences of HIV vulnerability, SRH, and the developmental, social, and institutional conditions within which biomedical HIV prevention is provided. By examining PrEP provision within the context of young women’s everyday lives, this study identifies the ethical considerations for supporting AGYW to access, engage with, and benefit from biomedical HIV prevention.

## 2. Method

### 2.1 Study design and setting

This qualitative study was nested within the Community-Based Chronic Care Lesotho (ComBaCaL) project, a population-based cohort evaluating community health worker-led models to strengthen primary healthcare delivery in rural Lesotho (Gerber et al., 2025). We explored how AGYW living in rural Lesotho understand and navigate SRH and HIV prevention within their everyday social and community contexts.

The study was conducted in the rural northeastern districts of Butha-Buthe and Mokhotlong. Each district comprises a central town surrounded by remote mountain villages situated at elevations of approximately 1,800–2,400 metres above sea level. Livelihoods depend largely on small-scale farming, informal employment, and seasonal labour migration to South Africa. Combined HIV prevalence across the two districts is estimated at 15.4% (Lesotho National Health Estimates, 2025). Some study villages were located near major infrastructure developments.

Many villages are geographically isolated, limiting access to healthcare, education, and other essential services. Each village is served by a community-elected Village Health Worker (VHW), who provides household-level care focused primarily on communicable and non-communicable diseases. At the time of data collection, VHWs did not routinely provide HIV prevention or SRH services. To capture variation in physical access to healthcare, villages were classified according to geographical accessibility based on whether travel to the nearest health facility required crossing a mountain or river, or walking more than 10 km (Gerber et al., 2025). These contextual characteristics provided an important setting in which to examine how social, institutional, and geographic conditions shape AGYW’s opportunities to engage with HIV prevention.

### 2.2 Participants & recruitment

Eligible participants were adolescent girls and young women (AGYW) aged 16–24 years who identified as female and lived in villages enrolled in the ComBaCaL cohort. Villages were selected using stratified purposive sampling by district and geographical accessibility. Within selected villages, we used purposive sampling to recruit AGYW in collaboration with VHWs (VHWs), who identified eligible participants through household records and village-level outreach.

A total of 93 AGYW were invited to participate across selected villages in Butha-Buthe and Mokhotlong. Thirteen focus group discussions (FGDs) were conducted across seven villages: five FGDs in three villages in Butha-Buthe and eight FGDs in four villages in Mokhotlong. Multiple FGDs were conducted in some villages to enable age-stratified discussions where feasible and accommodate participant availability. Groups included five to nine participants and were stratified by age where feasible (16–19 and 20–24 years) to reflect different developmental stages. Participants included school-going and out-of-school AGYW, as well as young women who were single, married, or parenting.

To complement AGYW perspectives, we conducted six interviews with community stakeholders, comprising two group interviews with VHWs and four individual interviews with village chiefs (VCs). Stakeholders were purposively selected because of their roles in community health and local leadership, providing insights into community norms, health service provision, and the broader social environment in which AGYW navigate SRH.

**Table 1.** Study sample.

| PARTICIPANT GROUP | TOTAL<br>(PARTICIPANTS, N) | BUTHA-BUTHE | MOKHOTLONG |
| --- | --- | --- | --- |
| ADOLESCENT GIRLS AND<br>YOUNG WOMEN (AGYW) | 12 FGDs (93) | 6 FGDs (52) | 6 FGDs (41) |
| **VILLAGE HEALTH WORKERS (VHWS)** | 2 FGDs (16) | 1 FGD (7) | 1 FGD (9) |
| **VILLAGE CHIEFS (VCS)** | 6 IDIs | 4 IDIs | 2 IDIs |

### 2.3 Data collection

Data collection took place between May and July 2024. One additional FGD was conducted as a pilot to refine the discussion guide and facilitation approach; pilot data were not included in the analysis. Separate semi-structured discussion guides were developed for AGYW and community stakeholders. The guides explored experiences of adolescence, SRH, HIV vulnerability, and HIV prevention, while allowing participants to introduce issues they considered important.

FGDs and stakeholder interviews were conducted in Sesotho by three trained research assistants (one female, two male) with prior experience conducting qualitative research in Lesotho. Each discussion was facilitated by one moderator and one co-facilitator responsible for note-taking and logistical support. Facilitators held bachelor’s degrees in public health, sociology, or related fields and received additional training in qualitative interviewing, research ethics, and adolescent-sensitive approaches before data collection. Facilitators had no prior relationship with participants.

FGDs and stakeholder interviews lasted approximately 60–90 minutes and were audio-recorded with participants’ consent. Facilitators kept field notes documenting non-verbal communication, contextual observations, and group dynamics. The research team held daily debriefing meetings to reflect on emerging findings, discuss field experiences, and inform subsequent data collection where appropriate.

### 2.4 Data analysis

We analysed the data using reflexive thematic analysis following Braun and Clarke (2006, 2019). Analysis was primarily inductive, with themes developed from participants’ accounts while remaining informed by the study objectives and discussion guide.

Audio recordings were transcribed verbatim in Sesotho and translated into English by bilingual members of the research team familiar with local dialects and terminology. A sample of transcripts was independently reviewed by a second translator to improve translation accuracy and minimise loss of meaning.

Analysis began alongside data collection, with insights from early discussions informing ongoing data collection through regular team debriefings. AW and TC familiarised themselves with the data through repeated reading of transcripts and field notes before independently coding the transcripts. Coding was conducted manually using printed transcripts and spreadsheets, and an example of the coding framework is provided in Supplementary Table 1.

Coding was iterative, with initial codes developed through close engagement with the data. Some codes reflected topics explored in the discussion guide, while others were generated inductively from participants’ accounts. Codes were reviewed, refined, and organised into candidate themes. Themes were developed through regular analytic discussions involving AW, TC, MM, LM, and CM, which challenged emerging interpretations, explored alternative explanations, and encouraged reflexive consideration of how researchers’ perspectives might shape interpretation while ensuring that themes remained grounded in participants’ accounts.

Participants did not review transcripts or findings because of the remote study setting, logistical constraints, and the group-based nature of the FGDs.

### 2.5 Researcher reflexivity

The study team included Basotho and international researchers with backgrounds in public health, epidemiology, and qualitative research. Data collection was conducted by trained Basotho research assistants who were familiar with the local language and sociocultural context but had no prior relationship with participants. The multidisciplinary analytic team brought experience in HIV prevention, adolescent health, and community-based research in Lesotho, while recognising that these professional and cultural perspectives inevitably shaped the interpretation of participants’ accounts.

Throughout data collection and analysis, the research team held regular debriefing and reflexive discussions to consider how their disciplinary backgrounds, cultural perspectives, and prior experiences might influence data collection and interpretation, consistent with a reflexive thematic analysis approach (Braun & Clarke, 2019). These discussions challenged emerging interpretations, explored alternative explanations, and supported the development of themes that remained grounded in participants’ accounts.

### 2.6 Ethical considerations

This study was conducted within the Community-Based Chronic Care Lesotho (ComBaCaL) cohort study (ClinicalTrials.gov: NCT05596773). Ethical approval for the ComBaCaL cohort was obtained from the National Health Research Ethics Committee of Lesotho (NH-REC; Ref: 210-2022) and the Ethics Committee of Northwestern and Central Switzerland (EKNZ; Ref: AO_2022-00058). Additional ethical approval for this qualitative study was obtained from the National Health Research Ethics Committee of Lesotho (Ref: 210- 2022NestedHIVPrev) and the University of KwaZulu-Natal Humanities and Social Sciences Research Ethics Committee (HSSREC; Ref: 0006690/2024).

All participants provided written informed consent before participation. For participants younger than 18 years, written parental or guardian consent and participant assent were obtained in accordance with national ethical guidelines. Participants were informed that participation was voluntary and that they could decline to answer any question or withdraw from the study at any time without consequence. FGDs and interviews were conducted in private locations to protect participants’ confidentiality. No identifying information was collected during discussions, and all transcripts were anonymised during transcription.

Electronic data were stored on password-protected servers and were accessible only to authorised members of the research team.

## 3. Results

Our findings describe deep intersectional vulnerability among AGYW in the context of a profound lack of social protection, with important implications for their sexual and reproductive health. Three interconnected themes emerged, illustrating how vulnerability develops during adolescence, deepens through early transitions into adulthood, and is reinforced by inadequate institutional responses.

### 3.1 Unprotected developmental transitions during adolescence

Participants described early adolescence as a period of limited access to SRH knowledge while simultaneously navigating early sexual relationships, age-disparate partnerships, and deep economic insecurity. These developmental transitions occurred with little guidance from parents, communities, or child protection structures during a life stage in which young women were still developing physically, emotionally, and socially.

#### 3.1.1 Limited sexual and reproductive health support

Open discussions about SRH were uncommon within households. Parents were frequently described as avoiding conversations about sex, leaving young women to navigate adolescence and emerging sexual relationships with little guidance:

“Some parents say they don’t talk to us about it (sex) because when they say ‘do not have sex’, you will do it anyway, it appears as if it’s the parents who taught you about it when they spoke to you about it.” - AGYW, Butha-Buthe

VHWs similarly recognised parents’ limited engagement in discussions about SRH, attributing this to parents’ own discomfort and lack of confidence in discussing these topics with their children:

"…as parents we don’t have relationships with our children. When they come to us about these issues we shut them out. As parents it’s like we aren’t equipped as to how we can approach our children. It is so difficult to sit them down, have a conversation with them about how this life works." – VHW, Mokhotlong

When conversations did occur, they were often only initiated after menarche and focused primarily on avoiding pregnancy rather than broader discussions about menstrual health, relationships, or sexual health. Many participants recalled being unprepared, describing the experience as confusing and distressing:

"At home when you first start to menstruate, you feel afraid, so afraid. Then your parents will start giving you a talk that boys, actually males will make you babies." - AGYW, Mokhotlong

Limited communication about SRH also constrained young women’s opportunities to engage with prevention services, illustrating that making services available did not necessarily mean they were accessible to young women:

"Even if we know where to access things like condoms, we are scared to go because of our parents. Even if we know we can get PrEP or PEP at the clinic, we can’t go because we are scared to ask for permission." - AGYW, Mokhotlong

Village chiefs and VHWs similarly recognised parental influence as an important barrier to young women’s engagement with SRH services:

"I also believe that it is not the issue of interest (in SRH services), rather the influence of parents." – VC, Butha-Buthe

"You will find that in some instances both parents and children don’t have the information, or sometimes the child has the knowledge about these issues but may be afraid to speak about going to get contraceptives because of what parents might say." – VHW, Mokhotlong

At the same time, young women described considerable social pressure to become sexually active while also experiencing persistent advances from older men. These relationships were frequently characterised as transactional and exploitative within contexts of poverty and economic vulnerability:

“Our friends will be telling us about what they do, how they are already doing sex and I will be pressured to engage in sex knowing exactly that I’m not ready.” - AGYW, Butha-Buthe

"They [men] do say a lot that when we are too old, our bones would have grown and hardened… these older men promise us money and also that we start doing sex while our bones are still soft." - AGYW, Butha-Buthe

Several participants suggested that the absence of parental guidance left young women more vulnerable to manipulation within unequal relationships:

"Our parents are still afraid to engage us in such things; as a result, we lack confidence and get manipulated by our boyfriends into unprotected sex." - AGYW, Butha-Buthe

#### 3.1.2 Sexual violence

Participants described experiencing multiple forms of sexual harassment, coercion, exploitation, and abuse across the spaces and institutions intended to support their development. Experiences of sexual violence occurred in the home, schools, hostels, and the vicinity of a large construction project in one district.

Some participants described experiencing sexual abuse within the household, including abuse perpetrated by a parent:

"Sometimes when I live only with a male parent. He would be forcing me to do sex." - AGYW, Mokhotlong

Rather than functioning as safe spaces for learning and development, schools were also places where young women experienced sexual harassment from those in positions of authority:

“Teachers also harass us sexually. They would be demanding us to do sex in exchange for marks.” - AGYW, Mokhotlong

Participants further identified hostels, where many students lived during the school term while attending secondary school, as important places of vulnerability. Older men reportedly waited near schools and hostels to target young women living away from parental supervision:

“Sometimes, you are a young girl and you are struggling to make ends meet and a stranger offers to take you to school and then that is where they trap you.” AGYW, Mokhotlong

A VHW similarly identified hostels as places where adolescents experienced increased freedom but limited supervision and support:

"Another challenge is that from age 13 upwards, is the age where children separate from parents due to going to secondary schools where they stay in hostels. That is where now they have too much freedom and where they have unplanned pregnancies and catching other infections… However, also if they had the knowledge perhaps we would be able to reduce unplanned pregnancies among young girls." – VHW, Mokhotlong

Participants also described the dam construction project in one district as creating new opportunities for the sexual exploitation of young women. Construction workers who had temporarily moved into the area were described as pursuing young women through relationships characterised by pronounced age and economic inequalities:

"We do sex at an early age because of poverty […] Older men working in construction along the roads do approach us." - AGYW, Mokhotlong

"It is really not easy for young people because these contractors ask them if they don’t trust them… Because this girl is just a child, she will agree knowing in their hearts that it’s not what she wants. But she will be coerced by Matala [an older man] because she also wants money." - AGYW, Mokhotlong

Some participants also described intentionally seeking relationships with construction workers because of their relative financial security:

"When we say we love money, the fact is that we date married men, especially contractors. They are the ones who are really popular because it is said that they pay money every month." - AGYW, Mokhotlong

Across both districts, relationships were frequently described as highly transactional and exploitative, with financial dependence limiting young women’s ability to refuse unwanted sex or leave abusive relationships:

“Sometimes they give us money without any relationships going on then later propose. And it becomes very difficult to refuse given the money you already received…. He is going to tell me I cannot refuse to have sex with him since he is giving me money." - AGYW, Mokhotlong

"He would tell me that he cannot raise a child for free, then manipulates me into unplanned sex. Then threatens me with death when I report. I would then keep quiet." - AGYW, Mokhotlong

### 3.2 Imposed early adulthood

Our findings describe how many young women assumed adult roles while still children. For many participants, the transition to adulthood was early and abrupt, commonly precipitated by early and unintended pregnancy and child marriage. Expectations associated with adulthood emerged before young women were developmentally, socially, or economically prepared to assume these roles.

#### 3.2.1 Unintended pregnancy and early marriage

Early and unintended pregnancy was common and often represented the beginning of an early transition into adulthood. As one village chief explained:

“Our biggest challenge is the children who get pregnant at a very young age. Indeed there is a high pregnancy rate in our village and early child marriage. And because they get pregnant, they find themselves getting married.” - VC, Butha-Buthe

Early pregnancy frequently led to early marriage, while early marriage itself carried expectations of childbearing, even in the absence of pregnancy. Pregnancy outside of marriage often resulted in young women being ostracised and losing support from their families, peers, partners, and the community.

“You will find out that when a friend gets pregnant, we are told to stop being friends because they will influence you to become pregnant also.” - AGYW, Mokhotlong

“‘You are now a woman, you can leave,’ (your family) say. For instance, if you need some bathing soap, they’d be telling you to go and ask for money from the father of the child, which was normally not the case.” - AGYW, Mokhotlong

"They (parents) start throwing mean words at you, then you end up deciding to get married to the father of your child." - AGYW, Mokhotlong

Responsibility for unintended pregnancy fell almost exclusively on young women, often in the absence of support from the father of the child. While young men continued their education and daily lives with few consequences, young women were expected to assume responsibility for childcare and caregiving.

“I was saying that he will go to school while he refuses to take responsibility and I’d be sitting at home with the baby.” - AGYW, Butha-Buthe

"As a result of unintended pregnancy, I am unable to go to school […] after falling pregnant, our male partners abandon us and avoid taking responsibility." - AGYW, Mokhotlong

Participants also described early and unintended pregnancy as disrupting educational trajectories through school absenteeism, social isolation, stigma, and exclusion from hostel accommodation.

“When you fall pregnant you have to stay out of school to avoid being judged and bullied, even the teachers look at you differently.” - AGYW, Butha-Buthe

"Sometimes if you get pregnant staying at the school boarding rooms, you are expelled out of your (hostel) room but still continue with school. Some drop out on their own without being expelled because they fear being shamed by their friends." - AGYW, Mokhotlong

In one district, participants described a deeply invasive practice of checking students for lactation to identify those who had previously been pregnant, referring to students "being milked":

"Sometimes, at school we know those who were pregnant and they will be called and their breasts will be milked. Usually, the principal calls for a meeting. Students and teachers will be called and the police will be informed. Then during this gathering each student will be milked." - AGYW, Mokhotlong

Despite assuming these adult responsibilities, participants repeatedly described themselves as emotionally young and longing for the freedom associated with childhood.

"If you are married at a young age, at some point you are still going to like the things of being a girl but it’s impossible now because you are now a married woman." - AGYW, Mokhotlong

"They [young girls] live in pain because this person still has a small mind, they will be scolded crying as if she is not a woman […] sometimes they wish to go out there to play just to freshen their mind but their husband would always be scolding her." - AGYW, Mokhotlong

Many participants described feeling unprepared for the responsibilities of marriage and reported returning home shortly after marrying.

"We don’t even last for a month in that marriage because we are still young, we end up returning from our marriages because we don’t know how to deal with marriage life and issues." - AGYW, Butha-Buthe

However, returning home was not always possible. In some communities, early marriage was described as an arrangement from which young women could not easily leave:

"Understanding is that it [early marriage] is marriage like any other. Even if parents do not like it, they cannot withdraw since it is a taboo for a girl to return from marriage." - AGYW, Mokhotlong

Participants also described profound emotional consequences of early pregnancy and child marriage. Across participant groups, unintended pregnancy, unsafe abortion, depression, and suicide were repeatedly identified as important concerns.

"Young people are engaging in sex at a young age even so, unprotected sex. So, some of them will even do unsafe abortions because their boyfriends would have refused the child." –VC, Mokhotlong

"This early child marriage can cause a lot of depression and a high rate of suicide." - AGYW, Butha-Buthe

"When someone finds out they are pregnant and they were not planning on having a child, they may end up thinking about committing suicide." – VHW, Butha-Buthe

### 3.3 Limited social protection and failures of the State

Our findings describe how the people and institutions expected to protect AGYW frequently failed to do so. Limited familial support, inadequate school responses, and weak community and institutional accountability left many young women to navigate violence, pregnancy, and early marriage with little protection or recourse.

Families are often the primary source of protection during adolescence. However, financial hardship, changing household circumstances, and abusive family environments frequently limited parents’ ability to fulfil this role:

"Our fields have been taken for building houses and camps. And when parents were compensated for the land, they spent the money on alcohol and looking out for themselves alone." - AGYW, Mokhotlong

"Parents encourage us to get married because they see certain opportunities in that family." - AGYW, Mokhotlong

One VHW identified abusive parental behaviour as a key driver of young women’s vulnerability:

"The type of abuse I see in my village is that of parents…let me put it this way, young girls end up running after older men because of the amount of abuse they receive from their parents in the family." – VHW, Mokhotlong

Similarly, other VHWs described parents themselves as struggling to fulfil their protective role under difficult social and economic circumstances:

"Often children end up in child marriages because we fail them as parents- we deviate from taking responsibility as parents. Sometimes, children end up married because they find peace with their boyfriends and not at home." – VHW, Mokhotlong

Participants also identified the large infrastructure project in one district as increasing young women’s vulnerability while describing little protection from its social consequences. One young participant reflected on the influence of the construction project on relationships between young women and construction workers:

"We are modern children, we are very unruly. Moreover, these construction workers there is no time that we are going to say they have abused us sexually because whatever they say we agree to, even if it’s your first time seeing them. Sometimes, you feel like you have feelings for them and end up having unprotected sex with them." - AGYW, Mokhotlong

A village chief similarly linked increasing vulnerability among young people to the construction project:

"We are affected by the Dam Construction… Children have become unruly due to this project." – VC, Mokhotlong

Schools also represented important institutions of support and protection during adolescence. However, despite national policies intended to protect pregnant learners, participants described schools that offered little practical support, leaving young women socially isolated and excluded from education:

"The teacher don’t even want to know who the father is. Once you fall pregnant you are on your own." - AGYW, Butha-Buthe

In some schools, exclusion extended to expulsion:

"Only the girl [gets expelled]. They say they can’t teach two people—you and your child." - AGYW, Butha-Buthe

Participants also described limited accountability when gender-based violence was reported to village chiefs or the police, with perpetrators frequently experiencing few consequences:

"The wife can report abuse to the chief, and the family will be called. Since the family is called, it always ends there with no consequences." - AGYW, Mokhotlong

"It doesn’t change anything even if you take him to the police. If there is nothing done to him, when he comes back he will still continue abusing you. But I would still report him." - AGYW, Mokhotlong

One VHW described a case in which community members actively assisted a perpetrator to evade accountability:

"I remember a case of an old man who impregnated an (adolescent girl), the VC was made aware and all the other responsible stakeholders were involved, even the police. There were people who were helping him escape." – VHW, Mokhotlong

## 4. Discussion

Our findings raise important ethical questions about the provision of PrEP to AGYW in in Lesotho in the absence of, or with insufficient attention to, the social protections required to support them. Adolescence is a uniquely vulnerable life stage, particularly for girls, during which rapid physical, emotional, and social development occurs and families, communities, and institutions play a critical role in supporting healthy development and reducing life stage-related vulnerability. However, for many participants, the transition into adulthood did not follow a typical developmental trajectory. Instead, early and unintended pregnancy and child marriage accelerated this transition, often before young women were developmentally, socially, or economically prepared to assume adult roles and responsibilities.

Our findings illustrate how an adolescent girl can suddenly become a wife, mother, daughter-in-law, caregiver, and sexual partner. With these roles, she is simultaneously conferred adult status and expected to assume adult responsibilities, often without the support that even many adults require. At the same time, the families, communities, and institutions expected to protect and support young women frequently failed to do so, with participants describing parental withdrawal, exclusion from school following pregnancy, and limited social and legal protection. We argue that the provision of PrEP to young women may implicitly carry a similar expectation of adulthood by assuming that they are prepared to independently assess HIV risk and take responsibility for protecting themselves from HIV, despite the broader social conditions that constrain their agency and increase their vulnerability. A strong case for strengthening social protection has recently been made in a population-based study conducted in Lesotho. The study found that social protection programs were associated with improved educational outcomes, increased consistent condom use, and reduced child marriage among adolescents living in poverty, highlighting the role of social protection in creating the conditions that enable healthier adolescent development and more effective HIV prevention (Hertzog et al., 2024).

Our findings are consistent with broader evidence demonstrating that early and unintended pregnancy and child marriage remain major challenges for AGYW in Lesotho. Nearly one fifth (19%) of girls aged 15–19 years have begun childbearing, with the highest rates occurring in the two districts where this study was conducted (Lesotho DHS, 2014).

Approximately one quarter of girls in Lesotho marry before the age of 18 years (Lesotho DHS, 2014). Across eastern and southern Africa, child marriage commonly occurs within contexts of poverty, early sexual debut, and transactional sexual relationships (Malhotra & Elnakib, 2021b; Siddiqi & Greene, 2022). Within these contexts, unequal gender and age power dynamics, limited SRH knowledge, and inadequate access to contraception further contribute to adolescent pregnancy.

Adolescent pregnancy is recognised as both a cause and consequence of child marriage, with poverty, family pressure, and stigma surrounding premarital pregnancy often contributing to early marriage—and vice versa (UNICEF, 2021; UNFPA, 2022). Consistent with this evidence, our findings illustrate how adolescent pregnancy, child marriage, and school dropout are closely intertwined. Previous research similarly shows that first births frequently precede marriage and that girls who leave school are more likely to marry young, while girls who marry young are more likely to discontinue their education (Petroni et al., 2017; UNICEF, 2021). Girls who marry before the age of 18 years are also at greater risk of sexual and intimate partner violence, while children born to mothers who married as children experience poorer health and social outcomes, perpetuating an intergenerational cycle of disadvantage (Kidman, 2017; Malhotra & Elnakib, 2021b).

In addition to the violence associated with child marriage, our findings provide insight into the multiple forms of sexual violence experienced by AGYW in Lesotho and highlight the need for greater attention to the role of sexual violence in shaping early sexual debut. Nationally, 11% of young women aged 18–24 years reported sexual debut before the age of 15 years, while nearly one fifth (18%) reported that their first sexual experience was forced or coerced (Violence Against Children and Youth Survey, 2018). A secondary analysis of the Violence Against Children and Youth Survey similarly found that although both girls and boys experience high levels of physical, sexual, and emotional violence before the age of 18 years, girls experience sexual violence three times more frequently than boys (15% versus 5%) (NICEF, 2023). Consistent with our findings, sexual violence occurred across multiple settings, including the home, community, and school, with girls reporting sexual violence perpetrated by a teacher.

Importantly, Lesotho has established several legal and institutional frameworks intended to protect AGYW, including the Children’s Welfare and Protection Act (2011), the School Health Policy (2005), the Prevention and Management of Learner Pregnancy Policy, and the Child and Gender Protection Unit (CGPU) of the Lesotho Mounted Police Service, which responds to domestic violence, gender-based violence, and child protection cases. However, our findings suggest that these protections are not consistently implemented. Participants described exclusion from school following pregnancy, limited accountability for perpetrators of sexual violence, and ineffective responses from community protection systems. Although the evidence base remains limited, one study similarly identified implementation challenges within the CGPU, including resource constraints, poor follow-up, and limited case resolution (Picchetti et al., 2022).

Our findings also identify important opportunities to strengthen existing social protection measures and develop new interventions. In particular, participants consistently identified school hostels as important sites where AGYW living away from parental support during the school term experienced heightened vulnerability. Participants also described how major infrastructure projects, such as the Polihali Dam project, created conditions that increased vulnerability to sexual exploitation, abuse, and, by extension, HIV acquisition. These findings support concerns previously raised by civil society organisations, which have documented increases in transactional sex associated with the project, alongside unintended pregnancies, school dropout, and unsafe abortions (Lesotho National Assembly, 2025).

Notably, these organisations have also argued that the Polihali Dam project commenced without adequately addressing longstanding community concerns regarding its social impacts, including risks to the sexual and reproductive health of AGYW.

## 5. Strengths and limitations

A key strength of this study was its exploration of the ethical dimensions of PrEP provision through the experiences of AGYW living in remote rural communities, complemented by the perspectives of village chiefs and VHWs who are embedded within these communities and work closely with young people. Although the findings are grounded in rural Lesotho, many of the challenges identified are consistent with evidence from other settings across eastern and southern Africa, suggesting that the ethical considerations highlighted in this study may be relevant in similar contexts. The study did not include the perspectives of parents, teachers, healthcare providers, or male partners, whose experiences may provide additional insight into the social, ethical, and practical considerations surrounding PrEP provision for adolescents.

## 6. Conclusion

Our findings call for a fundamental reconceptualisation of PrEP provision for AGYW in Africa, from a biomedical intervention originally developed for adult key populations to one designed for adolescents navigating development within highly gendered and socially constrained environments. Historically, PrEP has been framed as a response to individual HIV risk, implying that HIV prevention is primarily a matter of individual responsibility. Our findings instead suggest that HIV vulnerability is socially produced and requires equal attention to the developmental, social, and institutional conditions that shape young women’s ability to engage with and benefit from PrEP. Expanding access to PrEP is therefore necessary but insufficient. Ethical PrEP provision requires strengthening the social protections that enable AGYW not only to access biomedical HIV prevention, but also to meaningfully benefit from it.

## Author contributions

A. Williams contributed to conceptualisation, methodology, formal analysis, data curation, and writing the original draft. T. Crankshaw contributed to conceptualisation, methodology, formal analysis, writing – review and editing, and supervision. M. Strauss contributed to writing – review and editing and supervision. M. Motlalentoa, L. Mofilikoane, and M. Mohasoa contributed to investigation, data curation, and writing – review and editing. N. Labhardt contributed to conceptualisation, writing – review and editing, supervision, and funding acquisition. All authors read and approved the final manuscript.

## Acknowledgements

The authors would like to thank all the young women, village chiefs and VHWs who generously shared their time and experiences for this study. We also acknowledge the Lesotho Ministry of Health, the ComBaCaL team, and the local communities and district stakeholders in Butha-Buthe and Mokhotlong for their support with study implementation and data collection. Their contributions made this research possible.

## Data availability

The datasets generated and/or analysed during the current study are not publicly available because they contain potentially identifiable qualitative data. De-identified data may be made available from the corresponding author on reasonable request, subject to approval by the relevant ethics committees and in accordance with participant consent and applicable data-sharing agreements.

## Declaration of generative AI use

During the preparation of this manuscript, the authors used AI to assist with improving the clarity and readability of the text. The authors critically reviewed and edited all AI-generated content and accept full responsibility for the content of the published article.

## Abreviations

AGYW,: Adolescent girls and young women
ComBaCaL,: Community-based Chronic Care Lesotho
HIV,: Human immunodeficiency virus
PEP,: Post-exposure prophylaxis
PrEP,: Pre-exposure prophylaxis
SRH,: Sexual and reproductive health
VC,: Village Chief
VHW,: Village Health Worker

