## Supplementary Table 1 for "Biomedical Prevention without Social Protection: The ethics of providing PrEP to socially vulnerable adolescent girls and young women in rural Lesotho"

| **Initial code** | **Example quotation*** | **Analytic interpretation** | **Developing category** | **Final theme** |
| --- | --- | --- | --- | --- |
| Parents reluctant to discuss SRH | *"Some parents say they don't talk to us about it..."* | Limited parental communication constrained young women's preparation for sexual and reproductive health. | Gaps in developmental support | Unprotected developmental transitions during adolescence |
| Peer pressure to initiate sex | *"Our friends will be telling us about what they do..."* | Peer norms and social belonging influenced early sexual decision-making. | Peer-mediated transitions | Unprotected developmental transitions during adolescence |
| Transactional relationships with older men | *"...these older men promise us money..."* | Economic vulnerability reduced young women's ability to negotiate sexual relationships. | Transactional and age-disparate relationships | Unprotected developmental transitions during adolescence |
| Pregnancy leading to early marriage | *"...you end up deciding to get married to the father of your child."* | Pregnancy accelerated the transition into adult social roles and responsibilities. | Premature adult responsibilities | Imposed early adulthood |
| Unequal consequences of unintended pregnancy | *"...he will go to school while I'd be sitting at home with the baby."* | Responsibility for unintended pregnancy fell disproportionately on young women. | Gendered consequences | Imposed early adulthood |
| Sexual harassment by teachers | *"Teachers also harass us sexually..."* | Institutions intended to protect young women could also become sites of vulnerability. | Institutional failure | Limited social protection and failure of the State |
| Limited accountability following gender-based violence | *"...it always ends there with no consequences."* | Community and justice systems provided limited protection or recourse following violence. | Weak institutional response | Limited social protection and failure of the State |
| Fear of judgement at clinics | *"...we are scared to go because of our parents."* | Anticipated stigma and social judgement constrained engagement with HIV prevention services. | Health-service barriers | Limited social protection and failure of the State |
| Confidentiality concerns with local providers | *"...the nurses ask why young people like us need PrEP"* | Tensions between service accessibility and confidentiality influenced preferences for HIV prevention delivery. | Confidentiality and trust | Limited social protection and failure of the State |

**Supplementary Table 1. Illustrative example of the progression from initial codes to final themes during reflexive thematic analysis**

* *Illustrative excerpts only. Full quotations are presented in the Results*
